# The relationship between anxiety, health, and potential stressors among adults in the United States during the COVID-19 pandemic

**DOI:** 10.1101/2020.10.30.20221440

**Authors:** Angela M Parcesepe, McKaylee Robertson, Amanda Berry, Andrew Maroko, Rebecca Zimba, Christian Grov, Drew Westmoreland, Sarah Kulkarni, Madhura Rane, William Salgado-You, Chloe Mirzayi, Levi Waldron, Denis Nash

## Abstract

**Objective:** To estimate the prevalence of anxiety symptoms and the association between moderate or severe anxiety symptoms and health and potential stressors among adults in the U.S. during the COVID-19 pandemic

**Methods:** This analysis includes data from 5,250 adults in the Communities, Households and SARS/CoV-2 Epidemiology (CHASING) COVID Cohort Study surveyed in April 2020. Poisson models were used to estimate the association between moderate or severe anxiety symptoms and health and potential stressors among U.S. adults during the COVID-19 pandemic.

**Results:** Greater than one-third (35%) of participants reported moderate or severe anxiety symptoms. Having lost income due to COVID-19 (adjusted prevalence ratio [aPR] 1.27 (95% CI 1.16, 1.30), having recent COVID-like symptoms (aPR 1.17 (95% CI 1.05, 1,31), and having been previously diagnosed with depression (aPR 1.49, (95% CI 1.35, 1.64) were positively associated with anxiety symptoms.

**Conclusions:** Anxiety symptoms were common among adults in the U.S. during the COVID-19 pandemic. Strategies to screen and treat individuals at increased risk of anxiety, such as individuals experiencing financial hardship and individuals with prior diagnoses of depression, should be developed and implemented.

## Introduction

The novel 2019 coronavirus disease (i.e., COVID-19) pandemic has profoundly changed life in the United States (U.S.) in unprecedented ways. As of April 15, 2020, there over 570,000 confirmed cases of SARS-CoV-2 and 189,000 COVID-19-related deaths in the U.S.^1^ Although the COVID-19 pandemic remains unprecedented in our lifetimes, research conducted during and after the severe acute respiratory syndrome (SARS) epidemic in 2003 found that the prevalence of suicide, anxiety, and emotional distress increased during this period.^2-4^

Anxiety disorders are among the most prevalent mental health disorders in the U.S.^5^ Based on diagnostic interview data of a nationally representative sample of adults in the U.S., approximately 19% of adults in the U.S. had an anxiety disorder in the past 12 months and 31% had an anxiety disorder at some point in their lifetime.^5^ Anxiety disorders have been associated with poor physical health, co-morbid mental health disorders, and disability. Anxiety during the COVID-19 pandemic may be exacerbated by physical health concerns or ongoing stressors.

Individuals with certain underlying medical conditions including type 2 diabetes and obesity, are at increased risk of severe illness from SARS-CoV-2.^6^ Such individuals may experience greater anxiety during the COVID-19 pandemic. In addition, anxiety may be elevated among individuals with COVID-19-like symptoms or individuals who have recovered from COVID-19. Research during the SARS epidemic found that approximately half of individuals who had recovered from SARS experienced elevated levels of anxiety, and individuals who experienced severe illness during the SARS epidemic were at greater risk of subsequent depression and post-traumatic stress disorder.^3^

Anxiety during the COVID-19 pandemic may also be influenced by ongoing stressors, including financial or employment-related stress or concern for one’s physical health or the health of loved ones. The financial impact of the COVID-19 pandemic is vast and unparalleled in recent memory. As of April 2020, the U.S. unemployment rate was 14%, among the highest on record in the post-World War II era.^7^ Financial stress related to unemployment, underemployment, lost wages, or job insecurity may increase anxiety. For individuals employed during the COVID-19 pandemic, employment-related stressors may also contribute to increased anxiety. For example, healthcare workers may be at risk of anxiety due to increased workload, fatigue, inadequate personal protective equipment or other stressors.^8^ Other essential workers, including those employed in transportation, grocery stores, or pharmacies may experience anxiety related to concerns about COVID-19 risk and transmission, including inadequate personal protective equipment.

Greater understanding of the prevalence of and factors associated with anxiety symptoms during the COVID-19 pandemic is vital to inform the mental health response during and after the COVID-19 pandemic and identify populations particularly vulnerable to adverse mental health effects during the COVID-19 pandemic and future public health emergencies. This study aims to 1) estimate the prevalence of generalized anxiety symptoms among a geographically and socio-economically diverse sample of adults in the U.S. during the COVID-19 pandemic and 2) investigate the relationship between anxiety symptoms and health and potential stressors. This information can be used to guide the national and local mental health response to the COVID-19 pandemic throughout the U.S.

## Methods

### Sample

This analysis is based on the first round of recruitment and data collection of the Communities, Households, and SARS/CoV-2 Epidemiology (CHASING) COVID Cohort Study, a prospective cohort of US-based, geographically and socio-demographically diverse adults recruited during the COVID-19 pandemic in the U.S.^9^ Individuals who were 18 years or older and who resided in the U.S. or U.S. territories at enrollment were eligible for study enrollment. Study participants were recruited online via social media platforms or via referral. A total of 5,403 individuals completed the first study survey between April 3 and April 21, 2020. The current sample is comprised of the 5,250 individuals who completed this survey and for whom data on anxiety symptoms were available.

### Measures

#### COVID-related anxiety symptoms

COVID-related anxiety symptoms were measured using the Generalized Anxiety Disorder 7-item scale (GAD-7).^10^ The GAD-7 assesses the presence and severity of anxiety symptoms, with higher scores indicating greater severity. Participants were asked if they were bothered by any of the anxiety symptoms assessed “as a result of the coronavirus.” Anxiety symptom categories were defined as none (score, 0-4) mild (score, 5-9), moderate (score, 10-14) or severe (score ≥15). Binary classification of anxiety symptoms was defined as none/mild or moderate/severe.

### Health-related variables

#### Recent COVID-like symptoms

Participants were asked if they had cough, fever, or shortness of breath in the past two weeks. Participants who indicated that they had any of those symptoms were categorized as positive for COVID-like symptoms.

#### Daily smoking or e-cigarette use

Participants were asked the frequency with which they currently smoked cigarettes and the frequency with which they smoked electronic products (e.g., e-cigarettes, vape pens). Response options included not at all, some days, and every day. Individuals who responded that they smoked cigarettes or electronic products every day were categorized as daily smokers.

#### Depression

Participants were asked if a doctor, nurse, or other health professional had ever told them that they had depression. Individuals who responded yes were categorized as having a history of depression.

#### Underlying physical health condition

Participants were asked if a health professional had ever told them that they had: chronic lung disease, asthma (current), type 2 diabetes, a serious heart condition, kidney disease (not including kidney stones, bladder infection or incontinence), HIV, or immunosuppression. Participants who had been told they had any of those conditions were categorized as having an underlying physical health condition.

### Potential stressors

#### Health care worker

Participants were asked if in the past two weeks they had been involved in healthcare operations that involved diagnosis or care for persons who are confirmed or suspected to have COVID-19. This included people who delivered care and other services to sick persons, either directly as doctors, nurses, emergency responders, and home health care, or indirectly as hospital sanitation workers and medical waste handlers. Participants who indicated that they worked directly or indirectly in such health care operations were classified as health care workers.

#### Other essential workers

Participants were classified as other essential workers if they indicated that they worked in law enforcement, corrections or public safety, emergency management, delivery or pick-up services, or transportation.

#### Lost income in the past month due to COVID-19

Participants were asked if in the past month they had experienced a personal loss of income due to COVID-19. Individuals who responded yes were categorized as having lost income in the past month due to COVID-19.

#### COVID-related health worries

Participants were asked how worried they were about getting sick from COVID-19. Participants were also asked how worried they were about their loved ones getting sick from COVID-10. Answer choices included not at all worried, not too worried, somewhat worried, and very worried.

### Sociodemographics

Participants were asked to report their gender, age, race/ethnicity, income, employment status, and state and ZIP code of residence.

#### Rate of cumulative COVID-19 cases

Rates of cumulative COVID-19 cases (confirmed and probable) were assigned to each participant based on the county (or county equivalent) of residence and the date that the participant began the survey using data from The New York Times, based on reports from state and local health agencies.^11^ Crude rates were calculated using the total population from 2018 5-year American Community Survey estimates acquired via the National Historical Geographic Information System (NHGIS).^12^

#### City resident

Participants were assigned an indicator of “city resident” or “not city resident” based on their ZIP code of residence using data from the Education, Demographic, and Geographic Estimates (EDGE) program of the National Center for Education Statistics (NCES).^13^ If the participant’s home ZIP code, after conversion to ZIP Code Tabulation Area (a geographic unit which represents ZIP codes), was designated as being both an urbanized area and inside a principal city, they were assigned the “city resident” indicator.^13^

#### Geographic region of residence

Participants were assigned a geographic region of residence based on their reported state of residence at enrollment. Individuals who reported living in Alaska, Arizona, California, Colorado, Hawaii, Idaho, Montana, New Mexico, Nevada, Oregon, Utah, Washington, or Wyoming were designated as living in the West. Those who lived in Alabama, Arkansas, Florida, Georgia, Kentucky, Louisiana, Mississippi, North Carolina, South Carolina, Oklahoma, Tennessee, Texas, Virginia, or West Virginia were designated as living in the South. Those who lived in Connecticut, Washington, DC, Delaware, Massachusetts, Maryland, Maine, New Hampshire, New Jersey, New York, Pennsylvania, Rhode Island, or Vermont were designated as living in the Northeast. Individuals who reported that they lived in Puerto Rico or other U.S. territories were designated as living in U.S. territories. Individuals who reported living in Iowa, Illinois, Indiana, Kansas, Michigan, Minnesota, Missouri, North Dakota, South Dakota, Nebraska, Ohio, or Wisconsin were designated as living in the Midwest.

### Analysis

Univariate analyses were conducted to assess the prevalence of anxiety symptoms. Associations between key characteristics and severity of anxiety symptoms were analyzed using Pearson chi-squared tests. We used modified poisson regression to generate prevalence ratios for the relationship between anxiety symptoms and health and stressors.^14,15^ Adjusted analyses controlled a priori for age, gender, and race/ethnicity. For multivariable analyses, adjusted prevalence ratios (aPR) and 95% confidence intervals (CIs) are reported. This study was approved by the Institutional Review Board at the City University of New York Graduate School of Public Health.

## Results

Among the 5,250 participants surveyed, approximately one-quarter (27.0%) reported mild anxiety symptoms, 16.6% reported moderate anxiety symptoms, and 18.5% reported severe anxiety symptoms (Table 1). Approximately one-fifth (20.4%) of respondents were between 18 and 29 years of age and 27.2% were 60 years or older (Table 2). Just over half (50.2%) of the participants identified as female. Most (68.4%) participants identified as Non-Hispanic White. Age, gender, race/ethnicity, employment status, income, and having at least one child under age 18 in the home were all independently associated with the prevalence of moderate or severe anxiety symptoms among study participants.

**Table 1.**
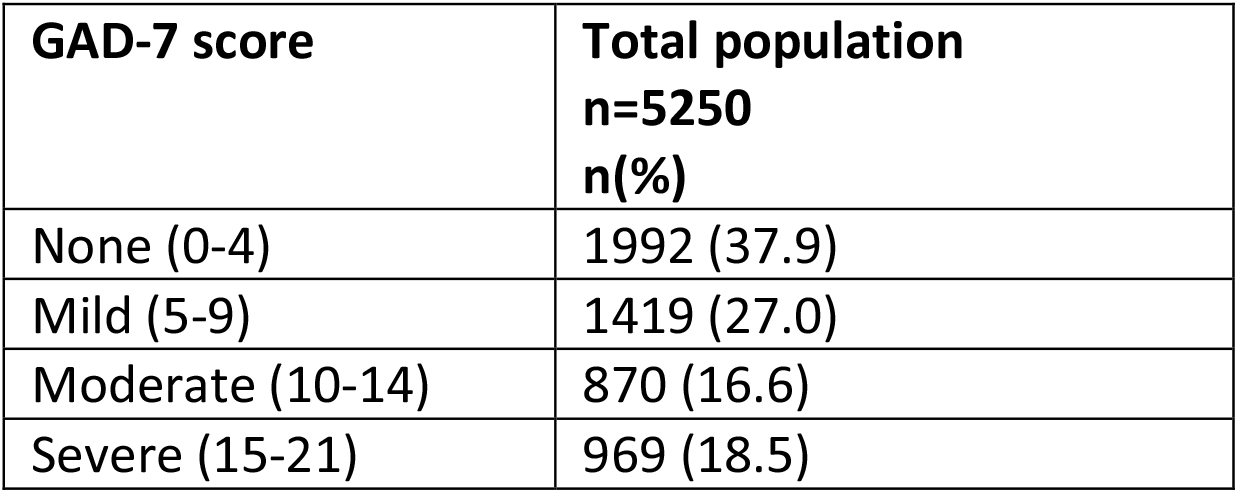
Generalized anxiety among adults in the U.S. during the COVID-19 pandemic.

**Table 2.** Sociodemographic Characteristics and Anxiety Symptoms among adults in the U.S. during the COVID-19 pandemic (n=5250)

|  | **Total sample population** | **None/Mild n=3411** | **Moderate/Severe n=1839** | **Chi square p-value** |
| --- | --- | --- | --- | --- |
|  | n (%) | n (%) | n (%) |  |
| **Age** |  |  |  | <0.0001 |
| 18-29 | 1070 (20.4) | 586 (17.2) | 484 (26.3) |  |
| 30-39 | 1007 (19.2) | 553 (16.2) | 454 (24.7) |  |
| 40-49 | 893 (17.0) | 532 (15.6) | 361 (19.6) |  |
| 50-59 | 851 (16.2) | 568 (16.6) | 283 (15.4) |  |
| 60+ | 1429 (27.2) | 1172 (34.4) | 257 (14.0) |  |
| **Gender** |  |  |  | <0.0001 |
| Male | 2461 (46.9) | 1737 (50.9) | 724 (39.4) |  |
| Female | 2633 (50.2) | 1610 (47.2) | 1023 (55.6) |  |
| Non-binary | 156 (3.0) | 64 (1.9) | 92 (5.0) |  |
| **Race/Ethnicity** |  |  |  | 0.003 |
| Hispanic | 674 (12.8) | 403 (11.8) | 271 (14.7) |  |
| Black/Non-Hispanic | 514 (9.8) | 350 (10.2) | 167 (9.1) |  |
| Asian/Pacific Islander | 251 (4.8) | 152 (4.4) | 99 (5.4) |  |
| White Non-Hispanic | 3593 (68.4) | 2377 (69.6) | 1216 (66.1) |  |
| Other | 218 (4.2) | 132 (3.9) | 86 (4.7) |  |
| **Employment** |  |  |  | <0.0001 |
| Employed | 2459 (46.8) | 1591 (46.6) | 868 (47.2) |  |
| Out of work | 927 (17.7) | 511 (15.0) | 416 (22.6) |  |
| Homemaker | 294 (5.6) | 177 (5.2) | 117 (6.4) |  |
| Student | 473 (9.0) | 248 (7.3) | 225 (12.2) |  |
| Retired | 1097 (20.9) | 884 (25.9) | 213 (11.6) |  |
| **Income (\$)** | | | | <0.0001 |
| <50,000 | 2674 (54.7) | 1629 (51.4) | 1045 (60.8) |  |
| 50,000 – 99,999 | 1210 (24.8) | 815 (25.7) | 395 (23.0) |  |
| 100,000+ | 1002 (20.5) | 724 (22.8) | 278 (16.2) |  |
| Missing | 364 |  |  |  |
| **Region** |  |  |  | 0.25 |
| Midwest | 1137 (21.7) | 755 (22.1) | 383 (20.8) |  |
| Northeast | 1227 (23.4) | 800 (23.5) | 427 (23.2) |  |
| South | 1780 (33.9) | 1130 (33.1) | 650 (35.4) |  |
| West | 1077 (20.5) | 704 (20.6) | 373 (20.3) |  |
| US Dependent | 29 (0.6) | 23 (0.7) | 6 (0.3) |  |
| **# children <18 living in household** |  |  |  | 0.0001 |
| 0 | 3920 (74.7) | 2604 (76.3) | 1316 (71.6) |  |
| 1+ | 1330 (25.3) | 807 (23.7) | 523 (28.4) |  |
| **Urban Area** | 1909 (36.4) | 1226 (35.9) | 683 (37.1) | 0.39 |

Underlying health conditions were common among study participants (Table 3). One-third (33.5%) reported an underlying physical health condition. Eighteen percent reported smoking or using e-cigarettes daily. Approximately 31% reported having received a diagnosis of depression. In bivariate analyses, all health conditions assessed were associated with the presence of moderate or severe anxiety symptoms.

**Table 3.** Health, potential stressors, and generalized anxiety among adults in the U.S. during the COVID-19 pandemic.

|  | **Total sample population** | **None/Mild**<br>**n=3411** | **Moderate/Severe**<br>**n=1839** | **Chi square p-value** |
| --- | --- | --- | --- | --- |
|  | n (%) | n (%) | n (%) |  |
| **Health-related factors** |  |  |  |  |
| **High Risk for Severe COVID19 Illness** | 2985 (56.9) | 1994 (58.5) | 991 (53.9) | 0.001 |
| **Any Underlying Health Condition** | 1756 (33.5) | 1087 (31.9) | 669 (36.4) | 0.001 |
| **Daily smoker** | 954 (18.2) | 518 (15.2) | 436 (23.7) | <0.0001 |
| **Depression Diagnosis** | 1635 (31.1) | 801 (23.5) | 834 (45.4) | <0.0001 |
| **Tested for COVID19** |  |  |  | <0.0001 |
| Yes | 202 (3.9) | 118 (3.5) | 84 (4.7) |  |
| No, but tried to be tested | 399 (7.7) | 211 (6.3) | 188 (10.4) |  |
| No, did not try to be tested | 4558 (88.4) | 3028 (90.2) | 1530 (84.9) |  |
| DK/Missing | 91 |  |  |  |
| **COVID-19-like symptoms past 2 weeks: cough, fever, or shortness of breath** | 943 (18.0) | 491 (14.4) | 452 (24.6) | <0.0001 |
| **All with COVID-like symptoms: cough, fever, or shortness of breath** | 943 | 491 | 452 |  |
| **Saw Physician** | 310 (33.1) | 157 (32.2) | 153 (34.0) | 0.55 |
| Missing | 5 |  |  |  |
| **Hospitalized** | 49 (5.2) | 25 (5.1) | 24 (5.3) | 0.88 |
| Missing | 4 |  |  |  |
| **COVID-19 Diagnosis (n=202)** | 67 (35.3) | 38 (33.6) | 29 (37.7) | 0.57 |
| Missing/Missing | 12 |  |  |  |
| **Potential stressors** |  |  |  |  |
| **Essential Workers** | 1185 (22.6) | 704 (20.6) | 481 (26.2) | <0.0001 |
| **All healthcare workers (HCW)** | 362 (6.9) | 207 (6.1) | 155 (8.4) | 0.001 |
| **HCW who screen or care for COVID patients** | 176 (3.4) | 100 (2.9) | 76 (4.1) | 0.02 |
| **Law enforcement** | 87 (1.7) | 56 (1.6) | 31 (1.7) | 0.90 |
| **Emergency response** | 51 (1.0) | 34 (1.0) | 17 (0.9) | 0.80 |
| Delivery services (e.g., food) | 590 (11.0) | 356 (10.4) | 234 (12.7) | 0.01 |
| Transportation (e.g., taxi, or airline) | 218 (4.1) | 121 (3.6) | 97 (5.3) | 0.003 |
| **Personal loss of income in past month due to COVID** | 2373 (45.2) | 1355 (39.7) | 1018 (55.4) | <0.0001 |
| **How worried are you about getting sick from coronavirus** |  |  |  | <0.0001 |
| Not at all worried | 481 (9.2) | 407 ((11.9) | 74 (4.0) |  |
| Not too worried | 1230 (23.4) | 1004 (29.4) | 226 (12.3) |  |
| Somewhat worried | 2323 (43.3) | 1522 (44.6) | 801 (43.6) |  |
| Very worried | 1216 (23.2) | 478 (14.0) | 738 (40.1) |  |
| **How worried are you about your loved ones getting sick from coronavirus** |  |  |  | <0.0001 |
| Not at all worried | 215 (4.1) | 185 (5.4) | 30 (1.6) |  |
| Not too worried | 551 (10.5) | 486 (14.2) | 65 (3.5) |  |
| Somewhat worried | 1809 (34.5) | 1410 (41.3) | 399 (21.7) |  |
| Very worried | 2675 (51.0) | 1330 (39.0) | 1345 (73.1) |  |
| **Rate of cumulative COVID-19 cases, County of residence (quartiles)** |  |  |  | 0.63 |
| Low ($\leq 37$) | 1294 (25.2) | 832 (25.0) | 462(25.6) | |
| Low-medium (38-69) | 1286 (25.1) | 822 (24.7) | 464 (25.7) |  |
| Medium-high (70-154) | 1273 (24.8) | 843 (25.3) | 430 (23.8) |  |
| High (155+) | 1278 (24.9) | 830 (25.0) | 448 (24.8) |  |
| Missing | 119 |  |  |  |

Potential stressors were also common among study participants (Table 3). In total, 22.6% of the participants were essential workers. Almost half (45.2%) reported a personal loss of income in the past month due to the COVID-19 pandemic, and 17.7% of participants were out of work at the time of the survey.

In multivariable analyses, having experienced COVID-19-like symptoms was associated with greater prevalence of anxiety (Table 4). The prevalence of moderate or severe anxiety symptoms among those with recent COVID-19-like symptoms was 1.17 (95% CI 1.05, 1.31) times the prevalence among those who did not report recent COVID-19-like symptoms, adjusted for age, gender, and race/ethnicity. Similarly, those who reported daily smoking had 1.17 (1.04, 1.31) times the prevalence of moderate or severe anxiety compared to those who did not report smoking daily. Having received a diagnosis of depression was associated with greater prevalence of moderate or severe anxiety symptoms (aPR 1.49 [95% CI 1.35, 1.64]) compared to not having received a diagnosis of depression. Having an underlying physical health condition was not meaningfully associated with the prevalence of moderate or severe anxiety symptoms in adjusted analyses (aPR 1.08 [95% CI 0.97, 1.19]).

**Table 4.** Associations among recent moderate or severe anxiety symptoms, health, and potential stressors among adults in the U.S. during the COVID-19 pandemic.

|  | **Moderate/Severe Anxiety Symptoms** |  |
| --- | --- | --- |
|  | **Bivariate**<br>**PR (95% CI)** | **Multivariable**<br>**aPR (95% CI)** |
| **COVID-like symptoms** |  |  |
| No | 1.00 | 1.00 |
| Yes | 1.49 (1.34, 1.66) | 1.17 (1.05, 1.31) |
| **Daily smoker** |  |  |
| No | 1.00 | 1.00 |
| Yes | 1.40 (1.26, 1.56) | 1.17 (1.04, 1.31) |
| **History of depression** |  |  |
| No | 1.00 | 1.00 |
| Yes | 1.83 (1.67, 2.01) | 1.49 (1.35, 1.64) |
| **Any underlying health condition** |  |  |
| No | 1.00 | 1.00 |
| Yes | 1.14 (1.03, 1.25) | 1.08 (0.97, 1.19) |
| **Loss income due to COVID (past month)** |  |  |
| No | 1.00 | 1.00 |
| Yes | 1.50 (1.37, 1.65) | 1.27 (1.16, 1.30) |
| **Worried about getting coronavirus** |  |  |
| Not at all/not too much | 1.00 | 1.00 |
| Some | 1.97 (1.72, 2.24) | 1.42 (1.22, 1.65) |
| Very | 3.36 (3.03, 3.96) | 2.00 (1.70, 2.36) |
| **Worried about loved ones getting coronavirus** |  |  |
| Not at all/not too much | 1.00 | 1.00 |
| Some | 1.78 (1.42, 2.22) | 1.45 (1.14, 1.84) |
| Very | 4.05 (3.29, 4.99) | 2.24 (1.75, 2.86) |
| **Essential health care worker** |  |  |
| No | 1.00 | 1.00 |
| Yes | 1.24 (1.05, 1.46) | 1.05 (0.89, 1.25) |
| **Other essential worker** |  |  |
| No | 1.00 | 1.00 |
| Yes | 1.16 (1.03, 1.31) | 1.09 (0.96, 1.23) |
| **Gender** |  |  |
| Male | 1.00 | 1.00 |
| Female | 1.32 (1.20, 1.45) | 1.16 (1.05, 1.29) |
| Trans/Non-binary | 2.00 (1.61, 2.49) | 1.26 (1.01, 1.58) |
| **Race** |  |  |
| Non-Hispanic White | 1.00 | 1.00 |
| Non-Hispanic Black | 0.96 (0.82, 1.13) | 0.94 (0.79, 1.11) |
| Hispanic | 1.19 (1.04, 1.36) | 0.97 (0.85, 1.12) |
| Asian/Pacific Islander | 1.16 (0.95, 1.43) | 0.99 (0.80, 1.22) |
| Other | 1.17 (0.94, 1.45) | 1.11 (0.88, 1.29) |
| **Age** |  |  |
| 18-29 | 1.05 (0.94, 1.18) | 1.14 (1.01, 1.28) |
| 30-49 | 1.00 | 1.00 |
| 50-59 | 0.77 (0.68, 0.89) | 0.82 (0.71, 0.94) |
| 60+ | 0.42 (0.36, 0.48) | 0.54 (0.46, 0.63) |
| **Urban area** |  |  |
| No | 1.00 | 1.00 |
| Yes | 1.03 (0.94, 1.14) | 1.01 (0.91, 1.12) |
| **Cumulative COVID-cases,**<br>**county of residence**<br>**(Quartiles)** |  |  |
| Low | 1.00 | 1.00 |
| Low-medium | 1.01 (0.89, 1.15) | 1.01 (0.89, 1.15) |
| Medium-high | 0.95 (0.83, 1.08) | 0.96 (0.84, 1.10) |
| High | 0.98 (0.86, 1.12) | 0.97 (0.85, 1.11) |

Potential stressors were also associated with anxiety symptoms among study participants (Table 4). Having lost income due to the COVID-19 pandemic was associated with greater prevalence of moderate or severe anxiety symptoms (aPR 1.27 [95% CI 1.16, 1.30]). Individuals who were somewhat worried (aPR 1.42 [95% CI 1.22, 1.65)] or very worried (aPR 2.00 [95% CI 1.70, 2.36]) about getting COVID-19 had greater prevalence of moderate or severe anxiety symptoms compared to those who reported being not at all or not too worried. Similarly, individuals who reported being somewhat worried (1.45 aOR [95% CI 1.14, 1.84)] or very worried (aOR 2.24 [95% CI 1.75, 2.86]) about their loved ones getting COVID-19 had greater prevalence of moderate or severe anxiety symptoms compared to those who reported not being not at all or not too worried. In bivariate regression analyses, being an essential health care worker and being a non-health care-related essential worker were associated with increased prevalence of moderate or severe anxiety symptoms. However, neither relationship persisted in multivariable analyses.

Individuals who identified as female (aPR 1.16 [95% CI 1.05, 1.29]) or transgender or non-binary (aPR 1.26 [95% CI 1.01, 1.58]) had greater prevalence of moderate or severe anxiety symptoms. There was a negative relationship between moderate or severe anxiety symptoms and age. Individuals between 18 and 29 years of age had significantly greater prevalence of moderate or severe anxiety symptoms (aPR 1.14 [95% CI 1.01, 1.28]) compared to those between 30 and 49 years of age. Those who were 50 to 59 years of age (aPR 0.82 [95% CI 0.71, 0.94]) and those who were 60 years of age or older (aPR 0.54 [95% CI 0.46, 0.63]) had significantly lower prevalence of moderate or severe anxiety symptoms compared to those who were between 30 and 49 years of age. In bivariate, but not multivariable regression analyses, identifying as Hispanic or Latino was associated with greater prevalence of moderate or severe anxiety symptoms. Neither living in a city nor the county-level cumulative COVID-19 case rate were meaningfully associated with moderate or severe anxiety symptoms.

## Discussion

Generalized anxiety symptoms were commonly reported among study participants, with over one-third (35%) of participants reporting moderate or severe anxiety symptoms. Several health-related factors, including smoking, depression, and recent COVID-19-like symptoms were associated with greater prevalence of moderate or severe anxiety symptoms. Potential stressors, including lost income due to the COVID-19 pandemic and worry about getting COVID-19, were also associated with greater prevalence of anxiety symptoms.

Prevalence estimates of anxiety among adults in the U.S. during the COVID-19 pandemic remain limited. However, one survey of adults in the U.S. during the COVID-19 pandemic estimated the prevalence of anxiety to be 25%.^16^ Caution is warranted when comparing estimates across studies due to differences in measurement and our non-random sampling strategy. A nationally representative survey of non-institutionalized adults in the U.S. conducted prior to the COVID-19 pandemic estimated that approximately 19% of adults in the U.S. had an anxiety disorder in the past 12 months.^5^ Our findings suggest that anxiety may be elevated among adults in the U.S. during the COVID-19 pandemic.

Smoking was associated with greater prevalence of moderate or severe anxiety symptoms. Prior research has found a consistently high co-occurrence of anxiety symptoms and smoking.^17,18^ Further, while smoking rates have declined in recent years among those without a mental health disorder, a similar decline in smoking has not been observed among individuals with an anxiety disorder.^19^ Understanding of the relationship between smoking and anxiety during the COVID-19 pandemic remains limited. One study conducted with adults in Australia during the COVID-19 pandemic found that the overwhelming majority of study participants did not smoke and reported no change in their smoking behavior since the beginning of the pandemic. However, among those who reported smoking prior to the pandemic, half reported an increase in smoking since the beginning of the pandemic.^20^ Further, the presence of anxiety symptoms was significantly associated with having increased smoking during the pandemic.^20^ Investigation of factors that increase one’s vulnerability to co-occurring smoking and anxiety, particularly during times of high stress and uncertainty, is needed. While evidence remains limited, early studies suggest that smoking may be associated with more severe COVID-related symptoms and worse health outcomes.^21^ Integrated interventions that address anxiety and smoking cessation and can be effectively delivered via telehealth may be particularly beneficial during the COVID-19 pandemic.

Having received a diagnosis of depression was also associated with greater prevalence of anxiety symptoms. Previous research demonstrates that anxiety and depression are highly comorbid.^22,23^ Transdiagnostic and cognitive-behavioral interventions tailored to address depression and anxiety offer promising approaches to effectively treat comorbid depression and anxiety.^24^ Such interventions have been successfully adapted for delivery over the internet, making them well suited to delivery during times of social distancing.^19^ Future research should examine the co-occurrence and trajectories of symptoms of depression and anxiety throughout the COVID-19 pandemic.

Having recently experienced COVID-19-like symptoms was associated with increased prevalence of anxiety symptoms. During the SARS epidemic, approximately half of individuals who had recovered from SARS reported symptoms of anxiety.^3^ Greater understanding of the mental health of individuals who have recovered from COVID-19 is urgently needed. Such research should investigate to what extent mental health symptoms are moderated by illness severity or length.

Worry about oneself or one’s loved ones getting COVID-19 was associated with anxiety symptoms. This is consistent with research with young adults in the U.S. that found high levels of COVID-19-specific worries associated with greater depression and anxiety.^25^ This research suggests that specific COVID-19-related worries may contribute to elevated levels of generalized anxiety during the pandemic. The extent to which strategies to manage COVID-19-specific anxiety influence generalized anxiety symptoms is worth investigation.

Financial stress, in the form of lost income, was significantly associated with anxiety symptoms. This is consistent with previous research that has found financial disruptions to be associated with worse mental health outcomes and poor quality of life. As the economic crisis in the U.S. worsens, the mental health of unemployed, underemployed, and furloughed workers warrants immediate attention. Uncertainty related to the continued provision of COVID-19-related financial relief may exacerbate anxiety symptoms. Research into factors, such as social support or coping strategies, that may modify the relationship between financial stress and anxiety should be investigated. Greater understanding is needed of the mental health impact of interventions that provide financial support and improve financial security.

Women and transgender or gender-nonconforming individuals reported a higher prevalence of moderate or severe anxiety symptoms. This is consistent with previous research that found the prevalence of anxiety to be significantly higher among women compared to men and among transgender and gender-nonconforming compared to cisgender individuals.^5,26^ Moderate or severe anxiety symptoms were also negatively correlated with age. Research conducted prior to COVID-19 found that older age groups experienced lower levels of anxiety.^5^ In addition, research conducted in the U.S. during the COVID-19 pandemic found anxiety symptoms to be more commonly reported among younger individuals. The current study found the highest levels of anxiety reported among the youngest participants. It is noteworthy that the negative relationship between age and anxiety persists during the COVID-19 pandemic, despite the fact that risk of severe COVID-19-related illness and COVID-19-related mortality increase with age. Younger populations may be more affected by direct and indirect economic consequences of the COVID-19 pandemic and may be more worried about the long-term impact of COVID-19 on employment, schooling, or financial trajectories.^27^ Additional research should examine factors that influence the prevalence of anxiety symptoms among younger populations during the COVID-19 pandemic.

Being an essential health care worker and being a non-health care-related essential worker were associated with prevalence of anxiety symptoms in bivariate, but not multivariable analyses. Previous research during the COVID-19 pandemic in the U.S. found that the prevalence of anxiety symptoms was meaningfully higher among essential workers.^10^ Differences in the categorization of essential workers and the measurement of anxiety symptoms across studies make direct comparisons challenging.

In bivariate, but not multivariable analyses, the prevalence of anxiety was greater among individuals who identified as Hispanic or Latino. Other research with adults in the U.S. during the COVID-19 pandemic found that those who identified as Hispanic had a higher prevalence of anxiety compared to those who identified as non-Hispanic Black, non-Hispanic White, or non-Hispanic Asian.^16^ Greater understanding of the mental health impact of the COVID-19 pandemic on communities of color and immigrant communities is urgently needed. The extent to which underlying structural inequities have been exacerbated during the pandemic and the impact on mental health warrants nuanced investigation.

This cross-sectional study has several limitations. The sample was recruited online or by referral and is not nationally representative of the adult population of the U.S. As such, findings cannot be generalized to the adult population of the U.S. Second, the directionality of relationships reported cannot be ascertained. In addition, all data were captured via self-report.

Anxiety symptoms appear to be elevated among adults in the U.S. during the COVID-19 pandemic. Longitudinal research to examine the persistence of anxiety symptoms among adults in the U.S. during and after the COVID-19 pandemic is needed. Such research can elucidate to what extent anxiety symptoms persist throughout different phases of the COVID-19 pandemic and its aftermath. Anxiety symptoms may diminish during the COVID-19 pandemic as individuals become more accustomed to social distancing measures. Alternately, anxiety symptoms may persist or worsen in response to national or local trajectories in COVID-19 cases, the easing or continuation of social distancing protocols, a worsening economic climate, or continued uncertainty. Strategies to screen and treat individuals at increased risk of anxiety, such as those experiencing financial hardship or with depression, should be developed and implemented. Evidence-based pharmacological and psychological interventions to treat anxiety, such as cognitive-behavioral therapy, should be implemented and scaled up for digital delivery throughout the U.S, particularly as remotely delivered psychotherapy interventions have been shown to be effective in treating anxiety.^28,29^ Medical professionals should consider routine anxiety screening and referral to care, as needed.

## Data Availability

The data that support the findings of this study are available upon reasonable request from CUNY ISPH. The data are not yet publicly available, but we are preparing to post a deidentified, HIPAA compliant, public use version of our baseline and follow-up data on GitHub.

## Acknowledgements

Funding for this project is provided by the CUNY Institute for Implementation Science in Population Health (cunyisph.org), the COVID-19 Grant Program of the CUNY Graduate School of Public Health and Health Policy, and the National Institute Of Allergy and Infectious Diseases of the National Institutes of Health under Award Number UH3AI133675 and NICHD grant P2C HD050924 (Carolina Population Center).

